# Exploring the use of generative AI advice for the academic advancement of faculty

**DOI:** 10.1101/2025.05.25.25328317

**Authors:** May May Yeo, Deborah D. Navedo, Patrick J. Casey, Bernard C.Y. Tan

**Author notes:** Corresponding author: (MMY).

## Abstract

The SingHealth Duke–NUS Academic Medical Center manages over 2,800 clinical faculty members and processes over 400 appointments and promotions annually. The current Promotion and Tenure documentation includes over 30 documents, making it difficult and time-consuming for the faculty to locate specific appointment information. We developed “AskADD” in response to requests for clearer academic career development guidance. This study reports initial alpha testing and subsequent beta testing with 35 faculty members using AskADD. AskADD aids the faculty—physician-educators, physician-scientists, physician-innovators, and physician-leaders—in navigating academic career paths while increasing transparency and trust in appointment, promotion, and tenure processes. Our AI-integrated systems, initially tested using low or no-code Microsoft platforms and later developed with a custom GPT, deliver contextualized responses to promotion and tenure queries. The faculty and staff participated in user testing, providing feedback for improvements. Alpha and beta testing conducted with the same group of users indicated that a substantial portion of participants found the tool beneficial; suggestions were given for further refinement. Our experience contributes to the limited literature on AI-driven faculty advancement in academic medical centers and offers a novel paradigm for academic career support.

## Introduction

The Singapore Health Services and Duke University-National University of Singapore Academic Medical Center (SingHealth Duke–NUS AMC) oversees more than 2,800 clinical faculty members and manages over 400 appointments and promotions annually. Promotion and tenure (P&T) information for the medical school is spread across 30 documents on the school’s intranet, which makes it time-consuming for the faculty to locate specific information on academic appointments. During engagement exercises with the school, the faculty expressed a need for more comprehensive guidance on academic career advancement. Our research team, which possesses expertise in institutional academic processes and artificial intelligence (AI), was uniquely positioned to address this problem. Combining our expertise, we were well-positioned to develop a tool to help the faculty navigate and explore academic careers. Our tool aimed to reduce the time spent by the faculty on searching for P&T information, increase transparency of the P&T policies and processes, and align faculty aspirations with the AMC’s mission and vision [1]. Using Microsoft’s Power Platform, including the Power Apps and Power Automate applications, and the OpenAI GPT-4o on Microsoft Azure (OpenAI GPT-4o, Redmond, WA, USA) [2,3] procured by the university, we developed an academic policy AI tool to help the faculty navigate P&T policies and processes. The tool could serve as a leadership development support resource for clinical program leadership and P&T committees, while reducing the administrative burden on staff by addressing common faculty queries through the chatbot. The faculty and staff were invited to test the tool and provide feedback before its launch, ensuring that it is effective and user-friendly.

Various studies have investigated the application of AI in vocational education career planning, AI mentors to navigate science, technology, engineering, and mathematics (STEM) careers; AI in shaping individual career paths across career stages, and AI in health profession education [4–9]. However, to our knowledge, no prior study has investigated the application of generative AI to support faculty candidates at AMCs in navigating policies and procedures related to promotion, tenure, and academic advancement. We aimed to address these gaps through the following two research questions—can a secure, interactive, online web application that allows faculty members to seek advice on academic advancement be developed, and how effective is generative AI in advising the faculty on academic advancement?

This study aimed to develop and test an academic policy AI tool to assist the faculty in navigating and understanding the policies and processes for academic appointments, promotions, and tenure at the SingHealth Duke–NUS AMC. We anticipate that when implemented, this tool will save time when searching for P&T policy information. Additionally, by providing contextualized natural language advice to faculty candidates preparing for promotion, our tool addresses their specific needs and increases their potential for success.

## Materials and methods

### Ethical approval

The National University of Singapore Institutional Review Board approved this study (date of approval: December 19, 2024; NUS-IRB Reference Code: NUS-IRB-2024-121). All participants were provided with an information sheet and completed a participant consent form.

### Study design, setting, and period

We performed alpha and beta testing and obtained feedback from participants on a newly developed AI chatbot to help the faculty navigate P&T policies and processes between January and April 2025.

### Alpha test

From August 2024 to January 2025, we programmed the university’s enterprise Microsoft Power Apps and Power Automate applications to contact the university’s enterprise OpenAI GPT-4o application programming interface and send user prompts to query a Microsoft SharePoint repository containing the school’s P&T policy documents. Responses from OpenAI GPT-4o could be viewed by the faculty through Power Apps application (Fig 1). Details of the tool, code and system prompts are provided in Supplementary Appendix S1 online.

**Fig 1.**
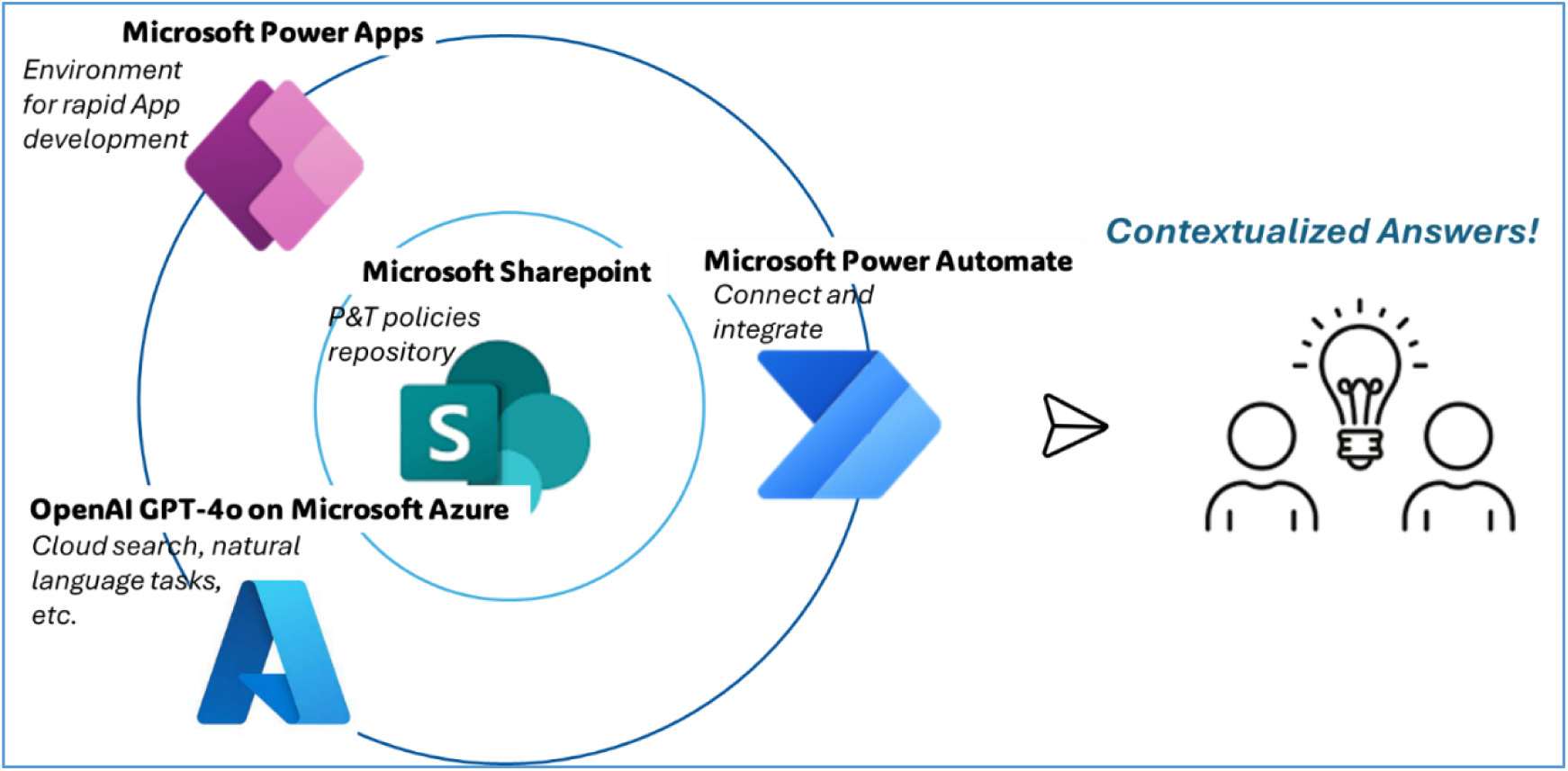
Using the university’s Enterprise Microsoft Power Platform and OpenAI GPT-4o on Microsoft Azure to generate natural language advice for the faculty. Microsoft Power Apps and Power Automate applications are programmed to contact Azure OpenAI GPT-4o to query Microsoft SharePoint repository containing policy documents. Responses from OpenAI GPT-4o can be viewed by users through Power Apps application.

### Study population, sample size and sampling procedures, data collection, and analysis

We launched the alpha version in mid-January 2025. To provide timely insights, we aimed to recruit a purposive sample of 30 participants [10]. We emailed 105 faculty and staff members of the SingHealth Duke–NUS AMC to participate. Non-faculty and administrative staff members of the hospitals were excluded from the invitation. In total, 36 faculty members and staff (34% response rate) agreed to participate. They represented a diverse range of user groups, including the faculty from various tracks, such as educator and research tracks, senior and junior faculty, clinical program leadership, and administrative staff who assisted the faculty in P&T processes. We obtained feedback from diverse user groups to help develop a tool that would be relevant and widely accepted [11].

Participants were requested to formulate queries, evaluate responses provided by the tool to them, and provide feedback on functionality and tool user interface via an online form (Supplementary Appendix S2) developed using Qualtrics (Provo, UT). We selected the following factors to assess chatbot user satisfaction: usability, which indicated the relevance and clarity of responses; responsiveness, which indicated the quickness and accessibility of services and perceived timeliness of responses; perceived empathy of the chatbot; and accessibility, wherein users with any browser had complete access to all the information and ability to interact with the chatbot. These factors were selected based on theoretical and empirical evidence supporting their relevance to AI chatbot interactions [12]. The Net Promoter Score [13], a measure of the participant’s likelihood of recommending the chatbot to colleagues, was added to complement the assessment by providing an action-focused indicator of user sentiment. Moreover, consultation with the university’s enterprise ‘NUS AI-Know’ informed the inclusion of an additional factor—comparison with traditional methods, such as administrative staff—to evaluate the chatbot’s perceived effectiveness as an alternative to human assistance [14]. Descriptive statistics were used to summarize participants’ feedback variables and plot data using Microsoft Power BI (Microsoft Corp.). Finally, we solicited additional feedback via two open-ended questions commonly used within the AMC’s faculty development programs: “What specific features of AskADD do you find most effective?” and “What improvements would you suggest in enhancing the effectiveness of AskADD?” Feedback was reviewed and interpreted with a reflective approach to capturing participants’ intended meanings, then summarized using a flexible thematic analysis framework [15]. Post-empirical or *a posteriori* themes or codes were developed progressively and added to a summary table. In developing the table, we noticed that the themes fitted into the factors of usability, responsiveness, accessibility, empathy, and comparative sources, and we termed them post-empirical or *a priori themes or codes*. Feedback related to two themes or codes was separated accordingly. Although feedback that did not align with existing themes or codes was initially intended to be categorized under “Other Comments,” no such feedback emerged during the alpha phase. Alpha testing concluded in the final week of February 2025.

### Alpha test feedback and custom generative pre-trained transformer (GPT) development in beta test

The university launched its enterprise “NUS AI-Know” platform in October 2024 and enabled the creation of custom GPTs which could be shared within its community in January 2025. Accordingly, we developed a custom GPT using NUS AI-Know. During the process, we incorporated participants’ feedback from the alpha test for improvement and adopted OpenAI’s prompt engineering strategies to enhance the results [16] (Supplementary Appendix S3). Beta testing was conducted from March to April 2025. The same group of faculty and staff was invited to submit queries, assess the tool’s responses, and provide feedback as previously described. All users except one responded, yielding 35 responses. To facilitate feedback comparison between both phases, we eliminated feedback from the alpha test that lacked corresponding beta test feedback.

In accordance with the university’s information technology data privacy policies, prompts in the tool were anonymized, and retained for auditing for 30 days. Users in beta were able to turn on “Private Mode” so conversations would not be saved. User prompts and usage statistics in both phases were not recorded by the research team. We launched the tool when we were satisfied with the accuracy of its responses and its functionality. Figure 2 summarizes the developmental steps.

**Fig 2.**
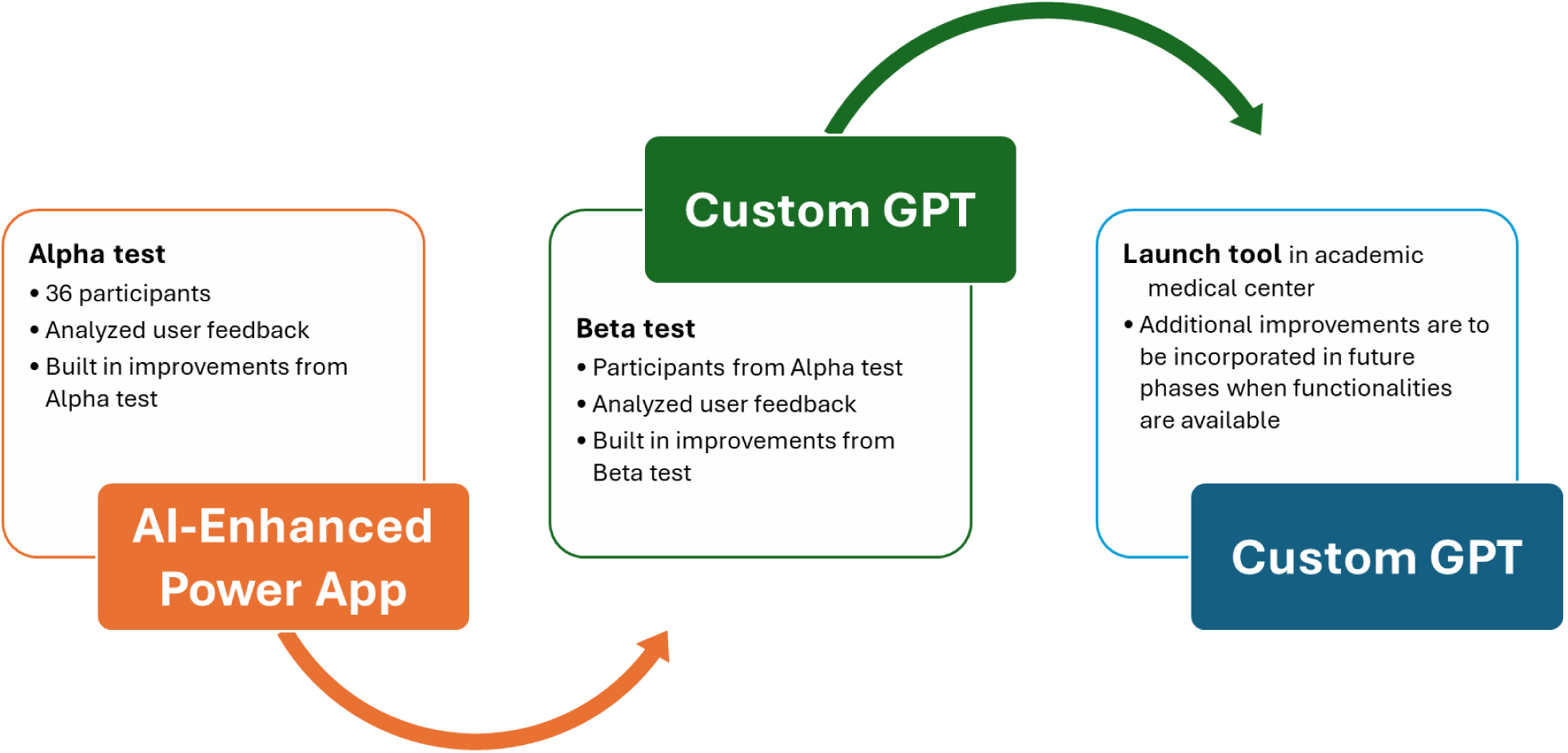
Steps in the development and implementation of a custom GPT to answer faculty questions on academic advancement. A flowchart illustrating an AI-integrated system, initially tested using low or no-code Microsoft applications and later developed with a custom GPT to deliver contextualized responses to users. This systematic approach ensures the GPT provides accurate and contextually relevant advice to support faculty advancement.

## Results

### Participant characteristics

There were 35 participants in both alpha and beta tests; more than half of them were men (57%). The participants predominantly comprised medical staff (60%), which corresponded with clinical faculty composition within our academic medical center. Furthermore, 40% of participants were in the clinical track, 26% were senior faculty, and 26% were engaged in school or institutional leadership (Table 1).

**Table 1.** Demographic characteristics of participants at SingHealth Duke-NUS Academic Medical Center (AMC) who participated in AskADD alpha and beta testing, 2025.

| *Category* | *Demographics (n=35)* | *Number* | *Percentage (%)* |
| --- | --- | --- | --- |
| **Professional group** | Administration | 5 | 14 |
|  | Allied Health Professional | 1 | 3 |
|  | Dental | 3 | 9 |
|  | Medical | 21 | 60 |
|  | Nurse | 1 | 3 |
|  | Research | 4 | 11 |
| **Faculty track**<br>*Where applicable* | Clinical | 14 | 40 |
|  | Educator | 5 | 14 |
|  | Practice | 1 | 3 |
|  | Research | 6 | 17 |
|  | Tenure | 4 | 11 |
|  | Not applicable, i.e., administrative | 5 | 14 |
| **Role** | Administrative Staff | 4 | 11 |
|  | Clinical Program Leadership | 8 | 23 |
|  | School or Institutional Leadership | 9 | 26 |
|  | Junior Faculty | 5 | 14 |
|  | Senior Faculty | 9 | 26 |
| **Sex** | Female | 15 | 43 |
|  | Male | 20 | 57 |

### Findings from alpha testing

Users felt AskADD provided relevant and clear responses and was effective in saving users’ time compared with searching the school’s intranet (Fig 3: Questions 1, 2 and 4 Alpha). Users also positively rated its responsiveness (Fig 3: Question 9 Alpha).

**Fig 3.**
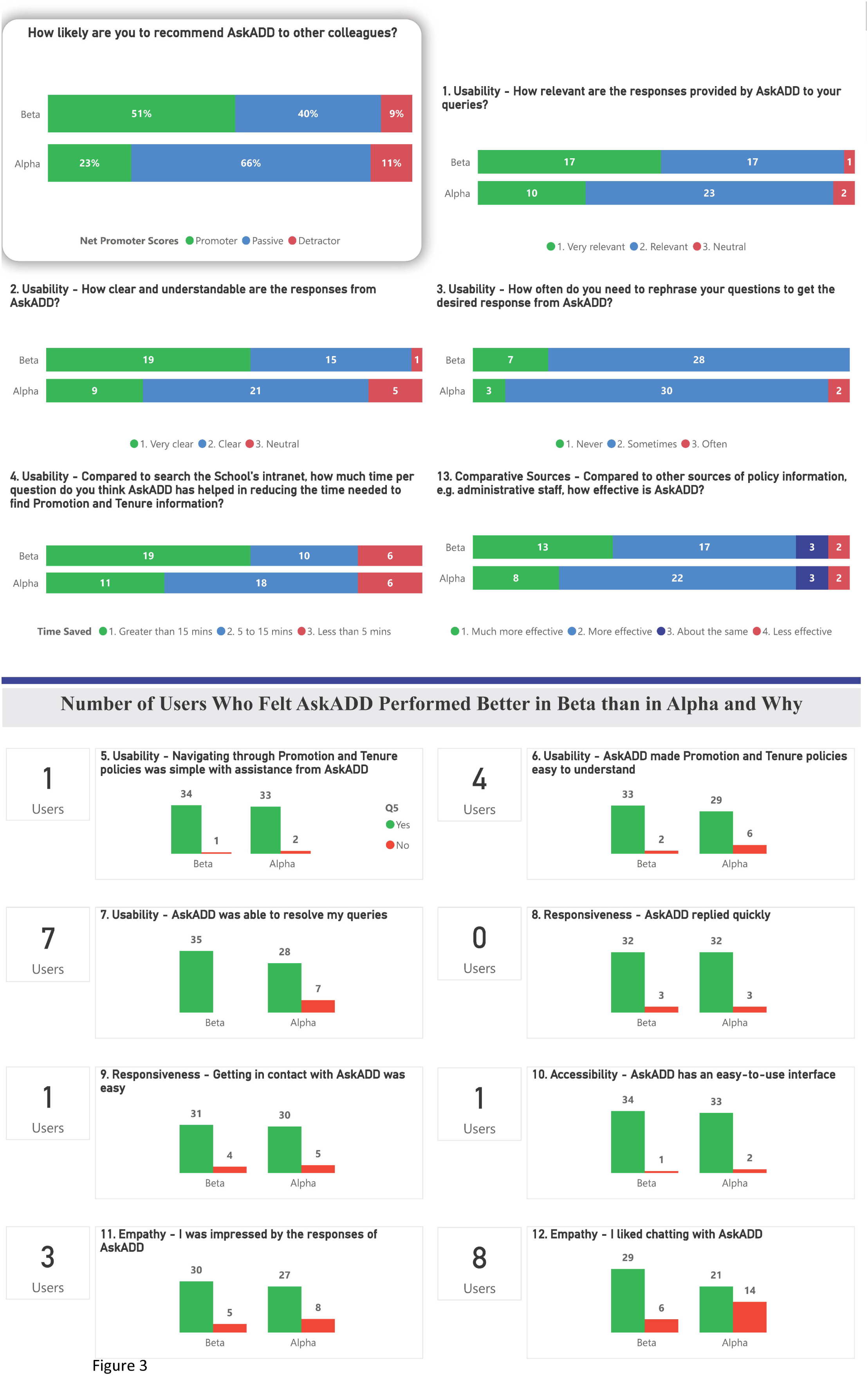
User feedback. User feedback comparisons denote overall improvement in the results of the beta over the alpha version.

32 users reported needing to rephrase queries, including 2 needing to rephrase often (Fig 3, Question 3 – Alpha). Between 23% and 40% indicated that there was room for improvement in the overall user experience (Fig 3, Questions 11 and 12 – Alpha). 66% of users rated the tool a 7 or 8 on a scale from 0 (not at all likely to recommend) to 10 (extremely likely to recommend) (Fig 3, Question “How likely are you to recommend AskADD to other colleagues?” – Alpha). These respondents were considered “passively satisfied” and indicated they would not actively recommend the tool to colleagues, as doing so might risk their professional credibility [11, p.4].

Additional feedback is presented in Appendix S4. Users felt that the chatbot would be improved by providing links to policy documents to enable users to verify information and explore further. It also had to improve handling of detailed follow-up advice, such as providing contact information to relevant departments. The chatbot should also include information on academic development and human resource information to accommodate a wider and related range of user needs.

### Findings from beta testing and comparison with alpha test results

Users reported that the custom GPT significantly reduced the time required to locate information. It was perceived as easy to interact with and responsive. Most participants reported that the tool was more effective than asking administrative staff for navigating academic policies. The Net Promoter Score increased from 12% in the alpha to 42% in the beta version.

### Usability

The proportion of participants who felt AskADD’s responses were very relevant increased from less than one-third (10 users) in alpha to approximately half of the participants (17 users) in the beta phase (Fig 3, Question 1). Similarly, the proportion of those who rated the responses as very clear rose from less than one-third (9 users) to over half of the participants (19, Fig 3, Question 2). The number of users who often had to rephrase their questions dropped to 0 (Fig 3, Question 3).

The proportion of users who reported that AskADD reduced the time required to locate the school’s P&T policies by more than 15 min increased from less than one-third (11) to over half (19, Fig 3, Question 4). However, the number of users who reported that AskADD reduced the time required to locate the school’s P&T policies by less than 5 min remained the same.

The number of participants who agreed that AskADD made it easier to navigate and understand policy documents increased from 33 to 34 and from 29 to 33, respectively (Fig. 3, Questions 5 and 6). All 35 users in beta phase felt that AskADD was able to resolve their queries (Fig. 3, Question 7).

### Responsiveness

Users in the beta reported similar turnaround times as those in the alpha phase (Fig. 3, Question 8). The number of users who found it easy to contact AskADD increased by 1 in beta phase to 31 (Fig. 3, Question 9).

### Accessibility

All except 1 user in the beta phase agreed that AskADD had an easy-to-use interface, up from 33 in the alpha phase (Fig. 3, Question 10).

### Empathy

Three additional participants were impressed by AskADD’s responses in the beta phase (Fig. 3, Question 11). The proportion of participants who enjoyed chatting with the tool increased from less than two-thirds (21 users) in the alpha to over 80% (29 users) in the beta phase. Those who did not enjoy the experience dropped to 6 in the beta phase (Fig. 3, Question 12).

### Comparing AskADD with other sources of policy information

The number of users who felt AskADD was much more effective than traditional sources increased by 5 to 13 in the beta phase (Fig. 3, Question 13). Although the majority considered AskADD a positive alternative for accessing academic policy information, about 15% or 5 users in both phases felt it was similar or less effective than other sources.

### Net Promoter Score and additional feedback

The proportion of promoters increased from 23% in the alpha phase to 51% in the beta phase. Passives decreased from 66% to 40%.

Figure 3 highlights two questions with the most notable improvement from alpha to beta: the tool’s ability to resolve queries (Fig. 3, Question 7) and perceived empathy (Fig. 3, Question 12), with gains of 7 and 8 users, respectively.

We explored the additional feedback to the two open-ended questions in the beta phase (Supplementary Appendix S4) to identify changes that could have contributed to the above outcomes. Features users found most effective included time-savings—*“Quick answers, don’t need to trawl website for info”*; relevant answers—*“Able to ask real questions and get directed in the right direction”*, hyperlinks to source documents: *“Excellent as is, especially updated version with links to the source references”*, and introduction prompts—*“Useful prompts to start a conversation”*. Users also requested streamlining access through the institutional network. There were also suggestions to provide document and application support, such as enabling users to upload documents for AskADD to provide personalized advice on academic rank and gap assessment. Where it could not provide an adequate answer, AskADD should be able to provide specific contact information. Users also suggested the appearance and speed of answers could be improved. The research team discussed and implemented the features where functionalities were available into the system before deployment.

### Additional findings from the tool development process

During the development of the alpha version, the team conducted approximately 60 iterations to enable hyperlinking to source documents stored in the SharePoint repository. In approximately 70% of cases, the Azure AI GPT-4o correctly returned file name formats, allowing the Power Automate application flow to retrieve and link the required documents seamlessly for user access. However, in the remaining 30% of runs, inconsistencies in the file name formatting generated by Azure AI GPT-4o disrupted the Power Automate application’s functionality—due to the latter’s reliance on standardized formatting—resulting in non-functional hyperlinks delivered via Power Apps. This limitation reduced the alpha version’s usefulness, which was addressed in the beta version through backend programming by the AI development team. Finally, on 20 iterations each of questioning both chatbots before launch, the alpha version required an average of 10 s, while beta was slightly faster at an average of 8 s. The team would explore using Azure AI GPT-4.1 launched in April 2025 when it was available to staff [17].

As accuracy and reliability of the tool were of primary importance for trust and acceptance in the chatbot [18], we took specific measures to reduce the risk of the chatbot generating incorrect information, a phenomenon known as “AI hallucination” [19]. Specifically, we largely focused on adjusting the temperature, a parameter that controls randomness of AI-generated responses to 0 and 0.3 in the alpha and beta versions, respectively [20]. A temperature of 0 produces consistent and predictable responses, while higher values (closer to 1) yield more variable and creative outputs [21]. While this safeguard did not eliminate the risk of misinterpretation, it significantly reduced instances of fabricated information. This ensured deterministic responses: when information was available, it was provided; when unavailable, AskADD clearly indicated that the requested information was not in its repository.

## Discussion

This study aimed to pilot an academic policy AI tool to parse P&T policies and to understand if it was effective in advising the faculty in an AMC on academic advancement. While we were able to create a tool using AI-enhanced Microsoft Power Platform in the alpha phase, the features and functionality needed improvement, several of which could not be implemented through the platform. Users requested the chatbot to improve its ability to interpret initial queries to reduce the need for rephrasing. Although the tool was perceived as effective compared to seeking human assistance, users required links to policy documents to verify and explore further and to provide contacts to dedicated support personnel if it was unable to answer queries. These requirements—to verify supporting documents and to reach dedicated personnel highlight the critical role of trust in acceptance and adoption of AI[22]. Users also required the scope of the tool to be broadened to include academic development pathways and human resources information. This enabled advice from a single source in place of searching in different chatbots for what users felt were related scopes – academic and human resource development. This finding adds to existing literature on professional development strategies to drive employees’ integration of AI to achieve organizational goals [23].

The custom GPT included several functionalities requested from the alpha tests: hyperlinks to policy documents, introduction to the chatbot and prompt questions to start conversations, improved system prompt guided by OpenAI, departmental contacts, and additional repository documents, including the AMC’s public websites to deliver holistic knowledge and answers to users. Thus, beta users felt the answers were relevant, clearer and required less querying effort. While the reported time saved was based on user perception rather than objective time-tracking with a control group, it nonetheless was an accurate reflection of the experiences of users that have likely had to obtain information via comparatively more time-consuming and likely tedious means, e.g., searching the school’s intranet.

The percentage of promoters more than doubled, with large improvements in the tool’s ability to resolve queries and its perceived empathy. Improvements in the system prompt strategy in the beta version likely contributed to a more engaging user experience and enhanced human–computer interaction. Nonetheless, the team noted that about half of beta users remained passives and detractors, pointing to the necessity of further improvements.

In relation to research question 2, users found the custom GPT helpful for addressing general queries, appreciating its round-the-clock availability in supporting their academic advancement preparation. However, users preferred personalized answers, such as if the tool could provide feedback and gap assessment upon evaluating their curriculum vitae and the academic level they intended to progress to. Such feedback resonates with discussions on chatbots as onboarding and career guides and providing real-time skills-gap analysis [24]. The team recorded the feedback for consideration and potential implementation in a future version.

## Limitations

Our study has several limitations. Firstly, we recruited a purposive sample of 35 volunteers from a single academic medical center. This volunteer-based approach may have introduced a selection bias, as participants were likely more digitally literate or motivated to engage with new technologies than the general faculty population, potentially inflating usability ratings. Furthermore, as the research team is affiliated with the institution, a social desirability bias may have influenced participants to overstate positive feedback to align with perceived expectations. We used results from the same group of 35 participants in both the alpha and beta testing phases. While this enabled direct comparison of user feedback between phases and strengthened the validity and credibility of the findings, familiarity with the tool and the research team may have influenced more positive results in beta.

Secondly, our methodology had inherent constraints. The primary outcomes, including time saved, usability, and empathy, were based on user perceptions rather than objective performance metrics. Consequently, inferences about the tool’s impact on career success are speculative, as we lack advancement outcomes. Additionally, as there were no “Other Comments” from the alpha phase, our feedback structure (framing versus predefined categories) may have constrained emergence of out-of-scope but relevant critiques.

Thirdly, the chatbot’s reliability is contingent on the currency and completeness of its underlying knowledge base. Therefore, outdated source documents risk generating incorrect advice and this necessitates a robust content management process. Furthermore, the risk of user over-reliance requires mitigation, such as disclaimers advising users to verify critical information with the original source documents. Moreover, the chatbot was intentionally limited to providing generic policy information. While some users desired personalized guidance, the tool’s lack of personalization is intended to circumvent issues tied to data privacy and security challenges inherent in handling sensitive employee data.

Finally, this study was based on platforms and tools that are available to and shareable among university staff. While this facilitates scalability within the institution, the chatbot uses Microsoft and university-specific infrastructure that may not be replicable in institutions lacking equivalent enterprise systems. Beyond the technical dependency on specific platforms, broader conceptual barriers to adoption exist at other sites. These include variations in institutional culture, differing levels of faculty trust in AI, and the significant variability of P&T policies across institutions, all of which can affect the tool’s implementation and acceptance elsewhere.

## Conclusion

We developed and demonstrated a secure, interactive tool that provided contextualized advice on institutional-specific P&T policies to faculty members. Our study revealed that users valued a chatbot that provides accurate, clear, and contextually relevant responses, with minimal need for query rephrasing. Users preferred seamless access to policy documents, direct contacts for unresolved queries, and a comprehensive platform that includes academic advancement and related human resource information in addition to P&T policies. While users valued a tool that could be accessed at all times, they felt the tool would be more effective if the answers were personalized through the input of personal information.

To the best of our knowledge, this is the first chatbot specifically designed to support faculty advancement at an AMC. Although tools, such as custom GPTs, are already procured for enterprise use at the university, we applied them innovatively to reduce the time faculty members spend searching for P&T information and to enhance their understanding of advancement pathways. Future studies should investigate the impact of user prompt formulation on response quality, as training users on effective prompting could improve the relevance and utility of the advice they receive. Other areas involve cautiously introducing personalized academic advisory features while navigating data privacy implications; exploring scalability across diverse institutional settings, and conducting longitudinal studies to evaluate the tool’s long-term impact on faculty advancement and satisfaction.

## Acknowledgments

The authors wish to thank the National University of Singapore Digital Enablement, Artificial Intelligence, and the Office of Data and Intelligence teams. We are grateful to Mr. Peng Wei, Mr. Tan Keok Tay, Mr. Manish Sridhar, Mr. Ku Wee Kiat, Mr. Irving Kwok, and Mr. Voycevestern Peh Jia Wei for their support and invaluable contributions. We also wish to thank Ms. Kelly Cheang and Ms. Tricia Wong of the Appointments and Development Department of Duke–NUS Medical School for their assistance with research.

## Author contributions

M.M.Y: Conceptualization, data curation, investigation, formal analysis, writing - original draft, review and editing. D.D.N. and P.J.C.: Formal analysis, writing - review and editing. B.C.Y.T.: Supervision, formal analysis, writing - review and editing, validation.

## Data availability

All data generated and analyzed during this study are included in this published article and its Supplementary information files.

## Supplementary information

Appendix S1

Appendix S2

Appendix S3

Appendix S4

