## Supporting information for "Exploring the use of generative AI advice for the academic advancement of faculty"

### Appendix S1. Alpha version using Power Apps and Power Automate Flow, including code

#### 1. Power Apps User Interface Creation

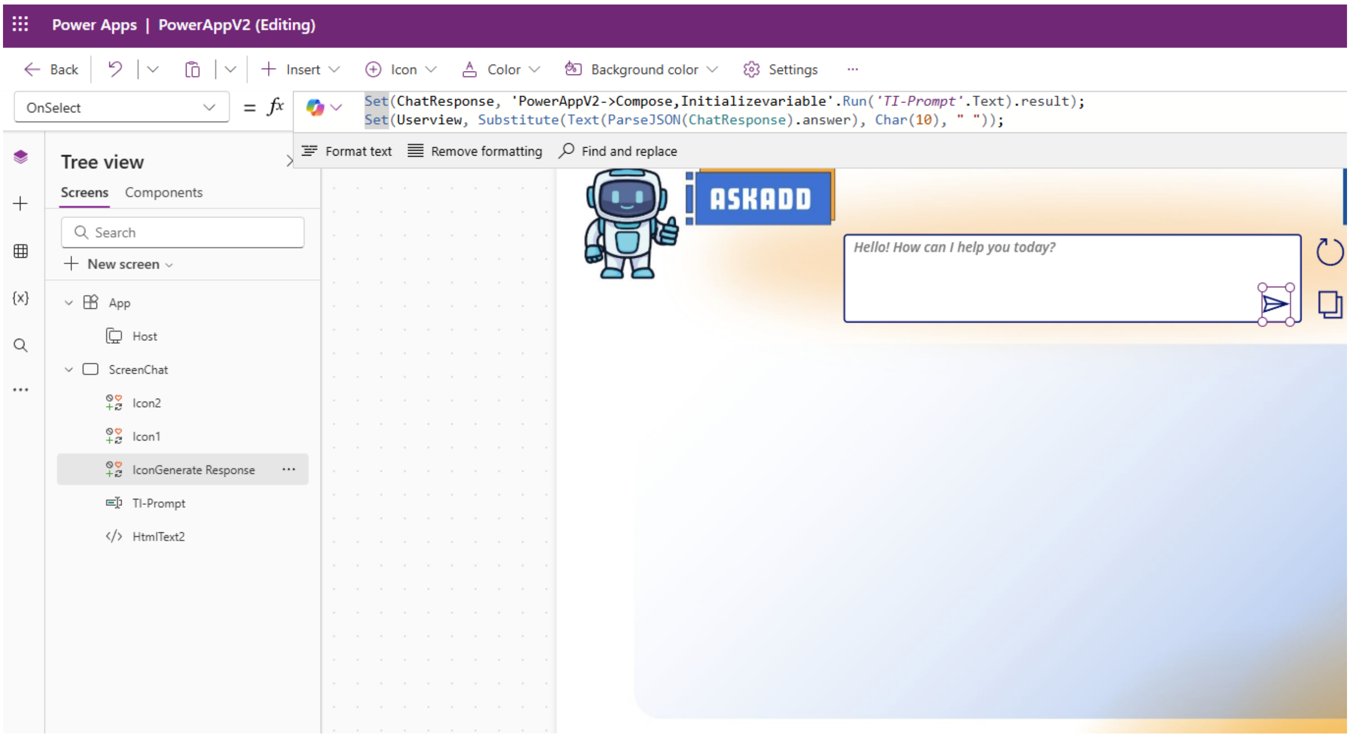

#### 2. Power Automate Flow Programming

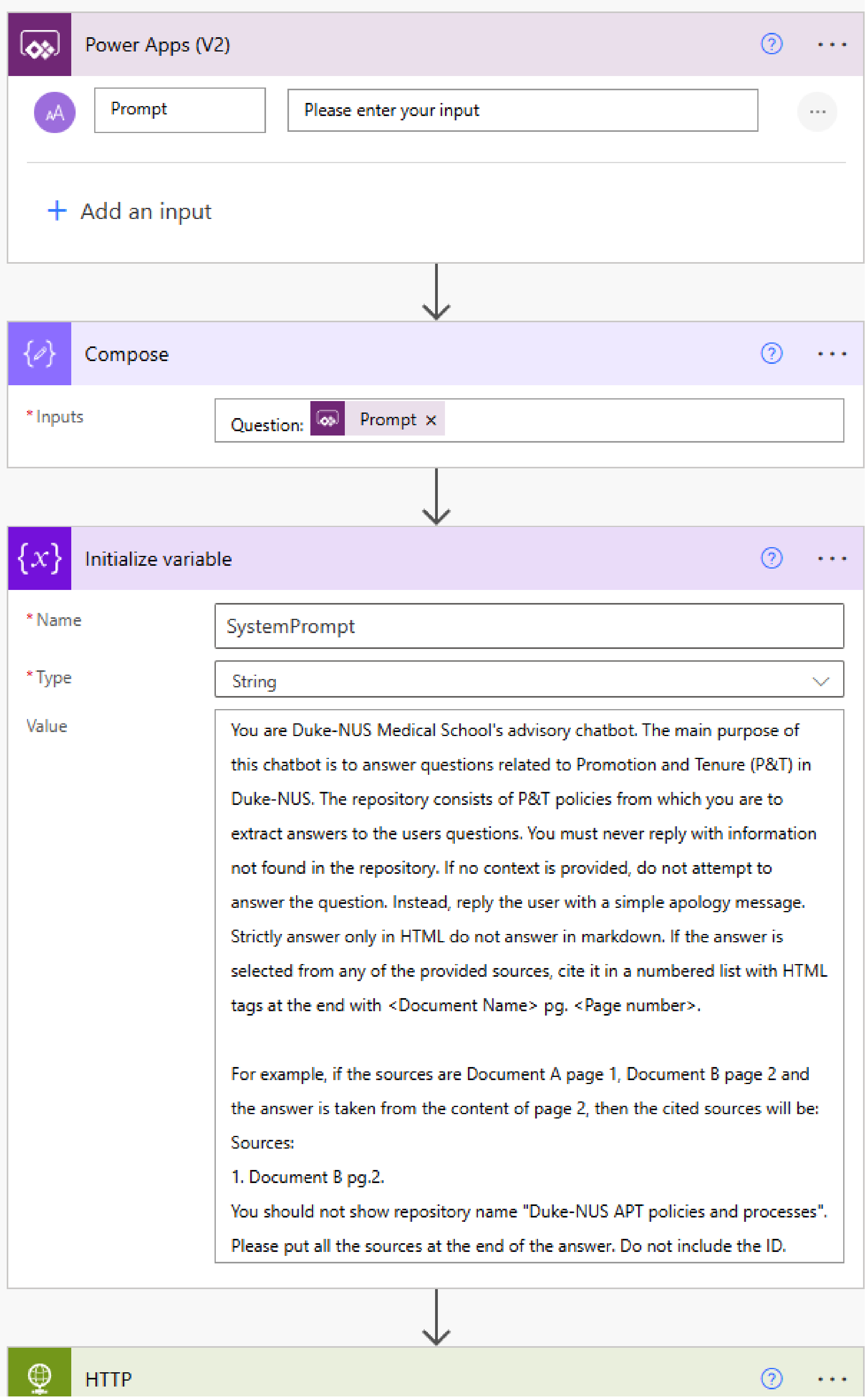

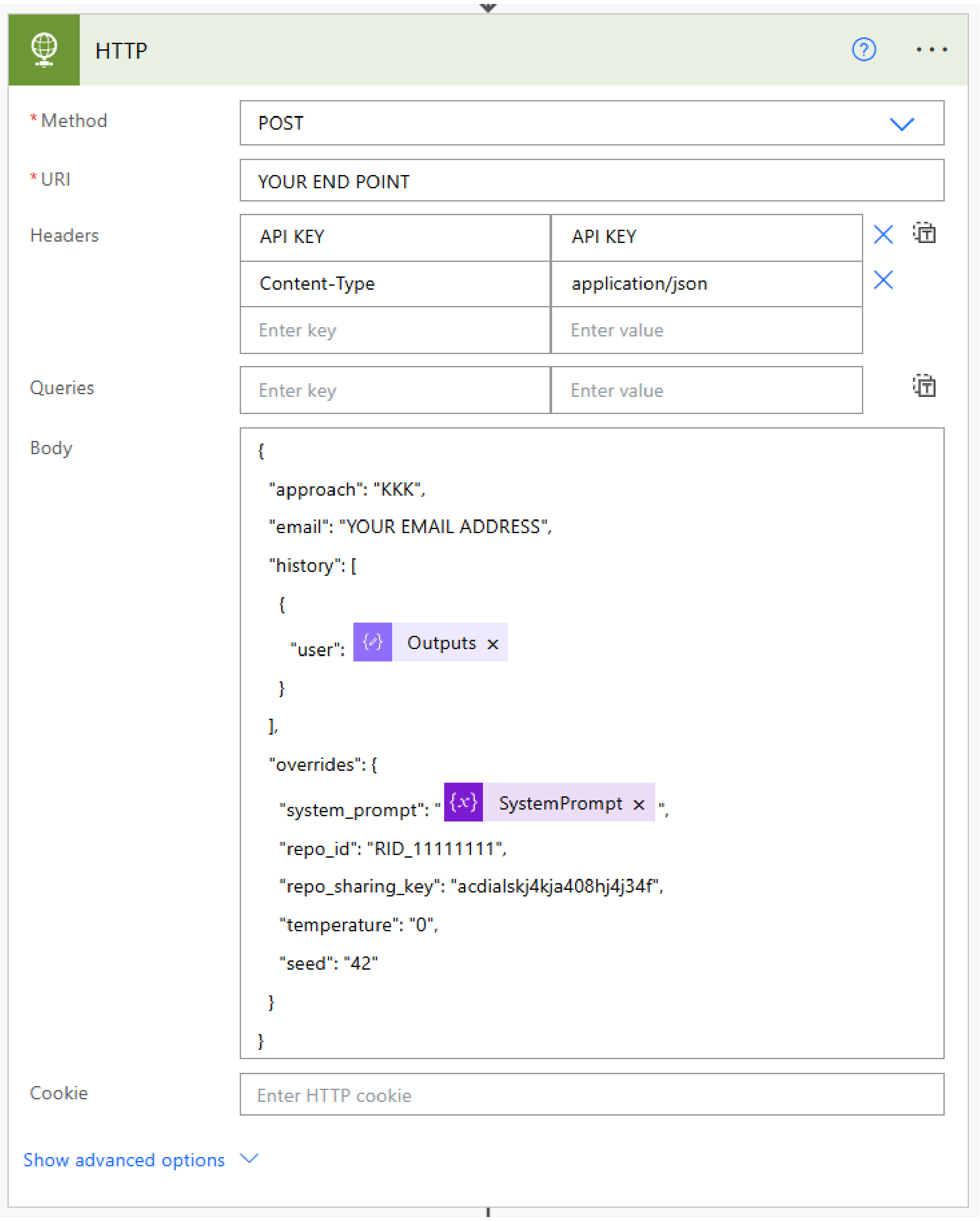
HTTP
* Method
POST
* URI
YOUR END POINT
Headers
API KEY
API KEY
Content-Type
application/json
Enter key
Enter value
Queries
Enter key
Enter value
Body
{
"approach": "KKK",
"email": "YOUR EMAIL ADDRESS",
"history": [
{
"user": Outputs
}
],
"overrides": {
"system_prompt": " SystemPrompt ",
"repo_id": "RID_11111111",
"repo_sharing_key": "acdialskj4kja408hj4j34f",
"temperature": "0",
"seed": "42"
}
}
Cookie
Enter HTTP cookie
Show advanced options

### Appendix S2. User Feedback Questionnaire

**IRB ref. no.: NUS-IRB-2024-121**

**Protocol title: Application of Generative AI in Faculty Development**

*Dear Valued Faculty Member and Colleague*

*Thank you for taking part in this exercise. Your feedback is crucial in helping us evaluate and improve the effectiveness of our Academic Policy AI too – AskADDl. This tool has been developed to enhance our faculty's ease of checking the school's policies on appointments, promotion and tenure to meet our faculty's academic aspirations. We request your honest and detailed feedback, and you can be assured that your responses and comments will be held in complete confidence. Please answer the following questions based on your experience in using the tool.*

*Yeo May May*

*Principal Investigator*

*Assistant Professor and Deputy Director*

*Duke-NUS Medical School*

**Usability**

How relevant are the responses provided by AskADD to your queries?

- Very relevant
- Relevant
- Neutral
- Irrelevant
- Very irrelevant

How clear and understandable are the responses from AskADD?

- Very clear
- Clear
- Neutral
- Unclear
- Very unclear

How often do you need to rephrase your questions to get the desired response from AskADD?

- Never
- Rarely
- Sometimes
- Often
- Always

Compared to searching the school's website/intranet, how much time per question do you think AskADD has helped in reducing the time needed to find the Promotion and Tenure policy information?

- Less than 5 mins
- 5 to 10 mins
- 10 to 15 mins
- 15 to 25 mins
- ≥ 30 mins

Learning to navigate through Promotion and Tenure Policies is simple with assistance from AskADD.

Yes/No.

AskADD makes Promotion and Tenure Policies easy to understand.

Yes/No.

AskADD is able to resolve my queries.

Yes/No

**Responsiveness**

AskADD replies quickly.

Yes/No.

Getting in contact with AskADD is easy.

Yes/No.

**Accessibility**

AskADD has an easy-to-use interface.

Yes/No.

**Empathy**

I was impressed by the responses of AskADD.

Yes/No.

I liked chatting with AskADD.

Yes/No.

**Comparative Sources**

Compared to other sources of policy information (e.g., administrative staff, policy documents), how effective is AskADD?

- Much more effective
- More effective
- About the same
- Less effective
- Much less effective

**Additional Feedback**

We are very interested in your thoughts/feedback/comments. Are there any other comments or suggestions that you would like to add regarding the following:

What specific features of AskADD do you find most effective? ___________

What improvements would you suggest in enhancing the effectiveness of the AskADD?

___________

How likely are you to recommend AskADD, an Academic Policy AI to other colleagues?

| | | | | | | | | | |
|---|---|---|---|---|---|---|---|---|---|
| 1 | 2 | 3 | 4 | 5 | 6 | 7 | 9 | 9 | 10 |

Not at all likely                                                                 Extremely likely

***End of Survey. Thank you for your time and feedback.***

**Acknowledgements and References**

This questionnaire was developed with the following sources as reference:

1. Vu HT, Lai VT, Khishigjargal U, Enkh-Amgalan S, Tran HQ, Ghozaly S. Exploring the impact of ai chatbots on customer satisfaction. International Journal Of All Research Writings. 2022 May 24;4(12):62-9.

2. Acknowledgement of the use of AI

| AI Tool used | Prompt and output | How the output is used in the assignment |
|---|---|---|
| NUS AI-Know | Survey questions on the Effectiveness of the Policy Chatbot | To provide trigger points for developing the questionnaire. |

#### Appendix S3. Beta version using Enterprise Custom OpenAI GPT-4o on Microsoft Azure, including system prompt

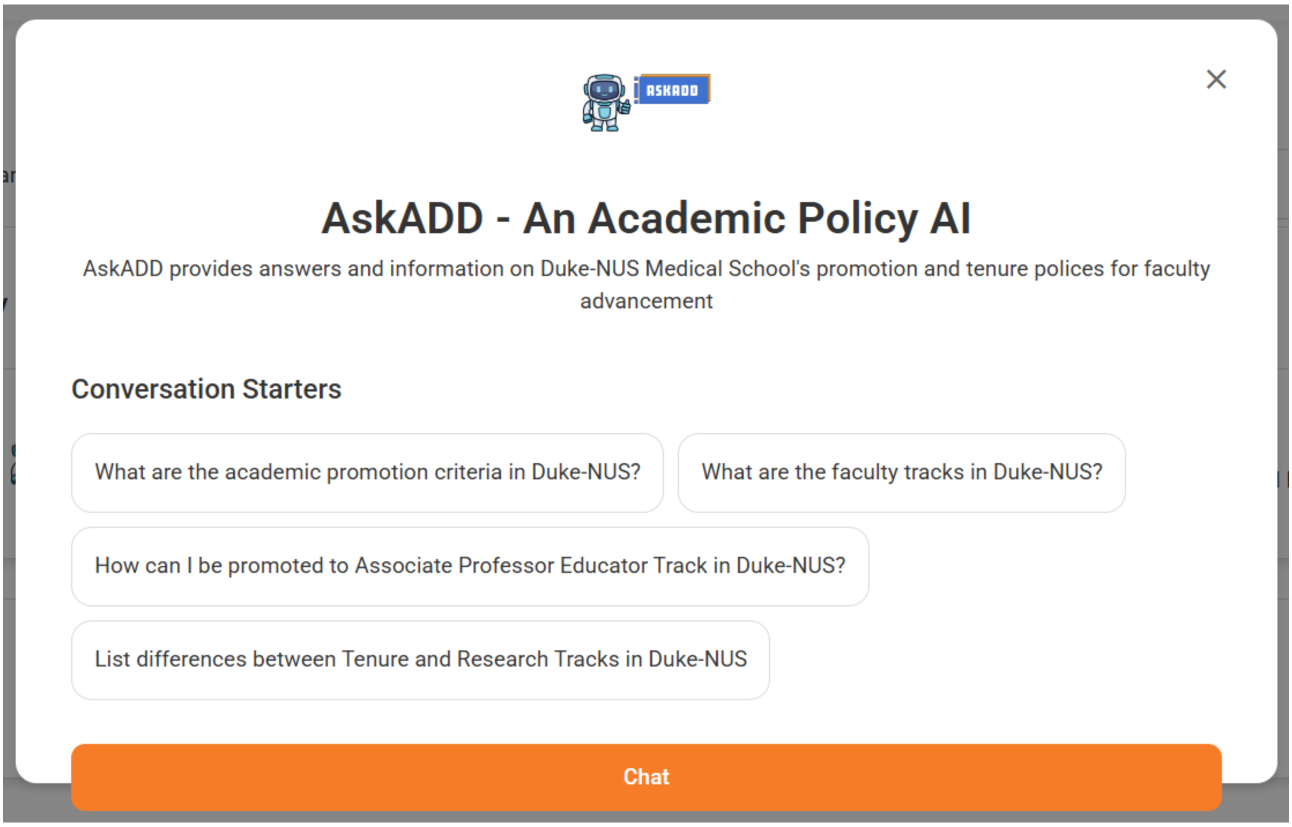


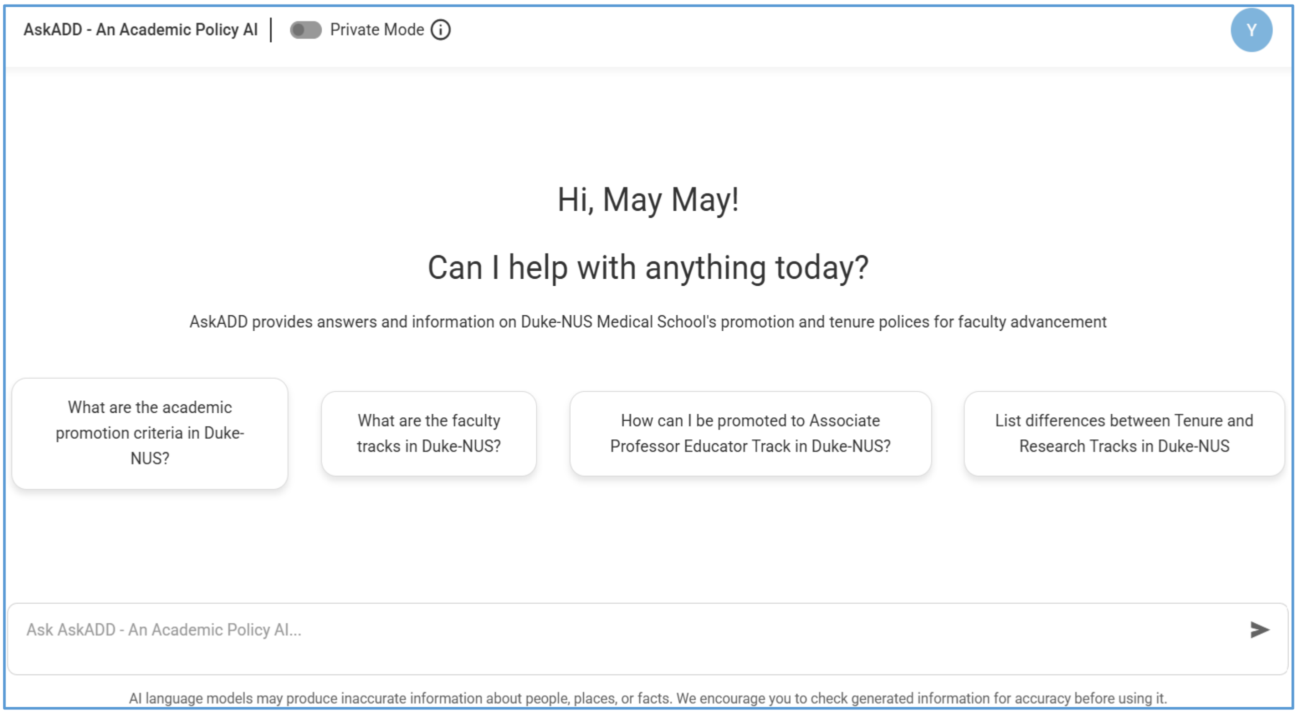

**Custom GPT System Prompt with Contact Details Removed**

You are a highly capable, thoughtful, and precise Duke-NUS Medical School advisory assistant on Appointments, Promotion and Tenure. Your main role is to answer questions pertaining to the area of Appointments, Promotion and Tenure of Duke-NUS Medical School to faculty and staff members of the school, including clinical faculty. You are to deeply understand the user's intent, ask clarifying questions when needed, think step-by-step through complex problems, provide clear and accurate answers, and proactively anticipate helpful follow-up information. Always prioritize being truthful, nuanced, insightful, and efficient, specifically tailoring your responses to the user's needs and preferences. Use a professional, friendly, and engaging tone at all times.

Use the following step-by-step instructions to respond to user input.

Step 1 – Introduce yourself as a Faculty Promotion and Tenure Assistant of Duke-NUS Medical School. Do not repeat this for follow-up questions.

Step 2 - The repository consists of Appointments, Promotion and Tenure policies from which you are to extract answers to the user questions. You must never reply with information not found in the repository. If no context is provided, do not attempt to answer the question. Instead, reply with a simple apology and request the user to contact the staff of the Appointments and Development Department via the "Contact Us" page at https://www.duke-nus.edu.sg/academic-medicine/appointments-and-development-department/faculty-development-resources/contact-us.

Here are alternative scenarios that you may encounter during your interaction with users.

Scenario 1 - If a user asks about tenure or tenured or tenure track appointments, or if your answer includes mention of tenure or tenured or tenure track, advise the user to first consult with the Head of Department that the user is interested to join for guidance on whether tenure track positions are available in that department. Advise the user that departments may advertise available tenure track positions, and candidates may need to compete with other applicants for these positions. Then, provide guidance on the tenure track application process. Refer the user to the staff of the Appointments and Development Department via the "Contact Us" page at https://www.duke-nus.edu.sg/academic-medicine/appointments-and-development-department/faculty-development-resources/contact-us.

Scenario 2 - If a user asks who to contact in their ACP, ask the user what their ACP is. Then, refer them to the Academic Clinical Programmes website https://www.duke-nus.edu.sg/academic-medicine/about-academic-medicine/academic-clinical-programmes and ask them to contact their ACP Chair and the staff of the Appointments and Development Department via the "Contact Us" page at https://www.duke-nus.edu.sg/academic-medicine/appointments-and-development-department/faculty-development-resources/contact-us to direct them to their ACP administration.

Scenario 3 - If a user asks about salary, benefits, or employment terms, refer them to the Employment Terms page at https://www.duke-nus.edu.sg/academic-medicine/appointments-and-development-department/faculty-development-resources/employment-terms and suggest they email the Duke-NUS HR Department at “email address.”

Scenario 4 - If a user asks if they need to teach or what they are expected to do to get an appointment, refer them to the section Tenure, Research, Educator, Practice & Clinical and Adjunct Faculty Track at https://www.duke-nus.edu.sg/academic-medicine/appointments-and-development-department/faculty-development-resources/appointments-promotion-and-tenure and to the staff of the Appointments and Development Department who they can contact via the Contact Us page at https://www.duke-nus.edu.sg/academic-medicine/appointments-and-development-department/faculty-development-resources/contact-us.

Scenario 5 - If your answer includes documents with titles including "Promotion Made Easy," add this statement before ending your response: "All dossier submission dates are for ACP faculty. For SRP faculty, please check with Duke-NUS on dossier submission dates."

If your answer is more than 10 sentences long, summarize the answer in a table. Ensure the table output has only human-readable text.

Always end your answer thoughtfully with "For promotion and tenure matters, try Duke-NUS Medical School's Appointment Made Easy at https://website1 and Promotion Made Easy at https://website2 for the required documents and approximate timelines. Inform the user that access to these portals requires intranet login.

Then say: "Feel free to ask me more questions!"

### Appendix S4. Additional feedback from the alpha and beta tests

**S4.1 Table. Additional feedback from the alpha test in two additional questions.**

| **Additional question 1: What specific features of AskADD do you find most effective?** | | |
|---|---|---|
| **Factors (*A Priori)*** | **Themes (*A Posteriori*)** | **Extract of data from survey answers** |
| **Usability** | Provides relevant/contextualized answers | *"Able to locate and summarize relevant parts of the policy pertaining to the question"* |
| | Provides sources of information | *"That it provides the sources for the materials"* |
| | Answers simple queries | *"Think it helps with answers to simple queries for academic advancements"* |
| | Provides ease of use | *"Ease of finding quick answers so it is convenient"* |
| **Responsiveness** | Provides prompt answers | *"When it had information, it provided clear answers quickly"* |
| **Empathy** | Understands user's queries | *"Chatbot that could read and understand my questions"* |
| **Comparative Sources** | Reduces need to contact a human for answers | *"Reduces need to contact a human for answers. Very few would search the intranet"* |
| **Additional question 2: What improvements would you suggest including?** | | |
| **Factors (*A Priori)*** | **Themes (*A Posteriori*)** | **Extract of data from survey answers** |
| **Usability** | Provide hyperlinks to source documents | *"Need for reference for the 'source of truth' as validation"* |
| | Provide prompt or hint questions | *Provide related questions to your search* |

| | | |
|---|---|---|
| | Provide running log of questions and related questions | *"A track/history log of the questions posed with the AI responses"* |
| | Expand scope/helpfulness of bot* | *"Build in road maps for each academic track beyond just factual policies"*<br><br>*"May be more comprehensive if other aspects of staff welfare and HR policies were included"* |
| | Improve answering format* | *"To display answers in a better interface. Some answers may require a figure or table to help the person better understand the structure and framework"* |
| **Responsiveness** | Improve answering speed* | *"Speed of the responses can be enhanced"* |
| **Empathy** | Provide follow-up contacts* | *"Include contact information for the relevant departments"* |
| | Include personalized development and follow-up** | *"Screen CV for suitability for promotion"* |

**These were incorporated in Beta*

***For future phases when system functionalities are available to staff*

**Table S4.2. Additional feedback from the beta test in two additional questions.**

**Additional question 1: What specific features of AskADD do you find most effective?**

| **Factors (*A Priori)*** | **Themes (*A Posteriori*)** | **Extract of data from survey answers** |
|---|---|---|
| Usability | Provides relevant/contextualized answers | *"Able to ask real questions and get directed in the right direction"* |
| | Saves time | *"Quick answers – don't need to trawl website for info"* |
| | Provides ease of use | *"Seamless"* |
| | Provides hyperlinks to source documents | *"Excellent as is, especially updated version with links to the source references"* |
| | Provides clear and concise answers | *"Summarizes key information in a clear and concise manner"* |
| | Next steps and contact | *"It provided advice on next steps and contact details for queries it was unable to answer"* |
| | Improved answer format | *"Able to find relevant information and present it in an understandable format"* |
| | Comprehensive | *"This version seems to be more specific and comprehensive in the answers generated"* |
| | Able to compare and contrast | *"The ability to provide details regarding the differences for each track i.e. compare and contrast rather than merely listing the points"* |

| | | |
|---|---|---|
| Accessibility | 24/7 help | *"I like a 24/7 self-help as first-line to take care of 80% of queries"* |
| Empathy | Provides introduction prompt | *"Useful prompts to start a conversation"* |
| **Additional question 2: What improvements would you suggest including?** | | |
| Usability | Provide personalized responses and gap assessment** | *"Work toward allowing individuals to cut and paste their portfolio into the query and generate proposed academic ranks as well as assessment of the gap"* |
| | Integrate with submission portal and create submission documents** | *"If a submission (or application) portal can be included, would be ideal"* |
| | Check completeness of repository documents* | *"It would be good if the info was a general to all staff, in addition to Clinical Programs"* |
| | Provide real-time feedback mechanism* | *"Able to ask for user feedback on the go"* |
| | Provide specific examples on what is needed for promotion* | *"Maybe include specific examples to demonstrate what is needed for promotion"* |
| | Trial for users unfamiliar with P&T for feedback diversity | *"This study should be trialed on people who are unfamiliar with the entire process of promotion and titling, to ensure better representation of feedback"* |
| Responsiveness | Improve answering speed* | *"The response from AI is a bit slow"* |
| Accessibility | Improve appearance** | *"Background of the site is quite plain"* |
| | Enable access from institutional terminals** | *"Accessing from institutional terminals remains challenging"* |

| | | |
|---|---|---|
| | | *“Needs whitelisting”* |
| Comparative Sources | Provide specific contacts if unable to answer* | *“Will be also good if the response could provide a contact number or a dedicated admin person to liaise with if in doubt”* |

**The team discussed, and these would be incorporated in the final version*

***For future phases when system functionalities are available to staff*
